# Integrating plasma, MRI, and cognitive biomarkers for personalized prediction of decline across cognitive domains

**DOI:** 10.1101/2024.12.31.24319830

**Authors:** Elaheh Moradi, Robert Dahnke, Vandad Imani, Christian Gaser, Alina Solomon, Jussi Tohka, the Alzheimer’s Disease Neuroimaging Initiative

## Abstract

**Background:** Plasma biomarkers are associated with cognitive performance and decline in Alzheimer’s disease, making them promising for early detection. This study investigates their predictive value, combined with non-invasive measures, for cognitive decline in non-demented individuals.

**Methods:** We developed a machine-learning approach incorporating plasma biomarkers (A***β***42/40, p-tau181, NfL), MRI, demographics, APOE4, and cognitive assessments. Various models were designed to predict decline rates across cognitive domains and assess their relevance in predicting dementia progression.

**Results:** Cross-validated correlations between predicted and actual cognitive decline rates were 0.50 for memory, 0.49 for language, 0.42 for executive function, and 0.44 for visuospatial ability. MRI showed greater predictive importance than plasma biomarkers. Among plasma biomarkers, NfL and p-tau181 outperformed A***β***42/40.

**Conclusion:** Plasma biomarkers, especially when combined with MRI, APOE4, and cognitive measures, have the potential to predict memory decline and assess conversion risk, even in cognitively unimpaired individuals.

## 1 Introduction

Alzheimer’s disease (AD) is a common neurodegenerative disease with a complex and unclear pathway and a long prodromal phase. Early detection and accurate prediction of AD are crucial for implementing timely interventions that may slow disease progression and improve patient outcomes. With several promising AD-modifying therapies currently in development [1, 2], the early identification of individuals at risk for cognitive decline has become increasingly important. As these therapies become available, accurately predicting cognitive decline at the patient level will be crucial for guiding clinical decisions. This ensures that costly treatments with potential side effects are administered only to those likely to benefit, thereby optimizing clinical outcomes and resource allocation.

Current diagnostic guidelines for AD involve detecting markers associated with brain amyloid-*β* (A*β*) plaques, aggregated tau (T), and neurodegeneration or neuronal injury (N) [3], typically measured using cerebrospinal fluid (CSF) [4] and positron emission tomography (PET) imaging [5]. However, these methods are often impractical for widespread use in community health settings due to their high costs and invasive nature. There is a critical need for more affordable, less resource-intensive, and widely accessible blood-based biomarkers [6]. Recent advancements in biomarker research have highlighted plasma biomarkers as a promising alternative. These non-invasive indicators of neurodegenerative processes facilitate earlier diagnosis and more effective monitoring of disease progression [7–10]. Key plasma biomarkers, such as A*β*42/40, p-tau 181, and neurofilament light (NfL), offer significant insights into AD pathology, aiding in a better understanding and earlier detection of the disease [8, 11].

While plasma biomarkers offer significant diagnostic accuracy for AD, they have limitations when used alone. The limited exchange of proteins between plasma and brain extracellular fluid can complicate the tracking of longitudinal changes in AD pathology [12, 13]. Integrating plasma biomarkers with other non-invasive measures, such as magnetic resonance imaging (MRI) and cognitive test results can enhance the predictive accuracy of AD conversion outcomes. This combination leverages the complementary strengths of various biomarkers, providing a more comprehensive understanding of the disease. Recent studies have underscored the importance of integrating different biomarkers to improve the predictive performance of early AD detection [14–17]. However, most previous research has focused on combining biomarkers such as MRI, cognitive test results, and PET imaging, with less emphasis on the role of plasma biomarkers in predicting cognitive decline across various cognitive domains [15, 18]. While recent studies have increasingly used plasma biomarkers to predict brain amyloidosis, their utility in predicting cognitive decline remains underexplored [10, 19].

This study aims to explore the role of plasma biomarkers in conjunction with other non-invasive biomarkers, including MRI data, demographic information, APOE4, and cognitive assessments, to predict the rate of cognitive decline in non-demented individuals using machine learning approaches. The specific objectives are to develop an ML-based approach by integrating different non-invasive biomarkers, including three key plasma biomarkers (A*β*42/40, p-tau 181, and NfL) to (a) predict the rate of cognitive decline across different domains, (b) investigate the relevance of various composite cognitive scores in predicting progression to dementia, and (c) predict progression to mild cognitive impairment (MCI) or dementia in cognitively unimpaired (CU) and MCI groups.

## 2 Materials and Methods

### 2.1 ADNI Data

Data used in this work were obtained from the ADNI (http://adni.loni.usc.edu). The ADNI was launched in 2003 as a public-private partnership, led by Principal Investigator Michael W. Weiner, MD. The primary goal has been to test whether serial MRI, PET, other biological markers, and clinical and neuropsychological assessment can be combined to measure the progression of MCI and early AD. For up-to-date information, see (www.adni-info.org).

This study included participants with baseline demographics, APOE4, plasma biomarkers (A*β*42/40, p-tau181, NFL), and MRI biomarkers, who also had available baseline and longitudinal composite cognitive scores. We included individuals who had at least one follow-up visit with available composite cognitive scores occurring two years or more after the baseline assessment. Since not all the selected participants had the plasma A*β*42/40 measure, we created two study cohorts for our analysis. Cohort 1 included participants with plasma biomarkers of NFL and p-tau181, but not A*β*42/40. The Cohort 2 included all three plasma biomarkers: A*β*42/40, p-tau181, and NFL. Cohort 2, which includes A*β*42/40, is a subset of Cohort 1. Having two cohorts is important because it allows us to utilize a larger sample size in Cohort 1, thereby improving the statistical power of our findings when A*β*42/40 is not considered. This approach is critical because NfL and p-tau 181 alone can still provide significant insights into cognitive decline and AD progression, as they are well-established markers of neurodegeneration and tau pathology, respectively [20, 21]. Cohort 2 allows us to specifically evaluate the added value of including the A*β*42/40 biomarker, which has been suggested to predict brain amyloidosis [22], a hallmark of AD. By conducting all experiments separately for both cohorts, we can compare the predictive power of models with different biomarker combinations, and assess the importance of A*β*42/40 in conjunction with other biomarkers. The baseline characteristics of study cohorts are presented in Table 1 and participant’s RIDs are available as supplementary material.

**Table 1.** Baseline characteristics of the study cohorts: The reported values for continuous measures are reported as mean(standard deviation). CN: cognitively normal, SMC: subjective memory concern (participants with self-reported significant memory concern), MCI: mild cognitive impairment, EMCI: early MCI, LMCI: late MCI. Classification of EMCI and LMCI is done by ADNI based on the WMS-R Logical Memory II Story A score. The specific cutoff scores were as follows (out of a maximum score of 25): EMCI was assigned for a score of 9-11 for 16 or more years of education, a score of 5-9 for 8-15 years of education, or a score of 3-6 for 0-7 years of education. LMIC was assigned for a score of *≤* 8 for 16 or more years of education, a score of *≤* 4 for 8-15 years of education, or a score of *≤* 2 for 0-7 years of education [23].

| Baseline characteristics | Cohort 1 (without Aβ42/40) | | | | Cohort 2 (with Aβ42/40) | | | |
| --- | --- | --- | --- | --- | --- | --- | --- | --- |
|  | CN | SMC | EMCI | LMCI | CN | SMC | EMCI | LMCI |
| Sample size (N) | 162 | 90 | 244 | 129 | 110 | 50 | 154 | 79 |
| ADNIGO/ADNI2 | 0/162 | 0/90 | 105/139 | 0/129 | 0/110 | 0/50 | 65/89 | 0/79 |
| Age, years | 73.3(6.4) | 72.1(5.7) | 71.1(7.3) | 71.6 (7.7) | 73.7(6.6) | 71.4(5.6) | 70.5(7.6) | 70.4(7.6) |
| Sex, M/F | 83/79 | 35/55 | 137/107 | 67/62 | 59/51 | 17/33 | 88/66 | 39/40 |
| Education, years | 16.7 (2.5) | 16.8(2.5) | 16.1(2.6) | 16.6(2.4) | 16.7(2.6) | 16.3(2.4) | 16.1(2.7) | 17.0(2.3) |
| APOE4 (0/1/2) | 117/40/5 | 59/30/1 | 142/84/18 | 59/52/18 | 79/28/3 | 33/16/1 | 95/51/11 | 36/30/13 |
| ADNI-MEM | 1.07 (0.6) | 1.07(0.57) | 0.62(0.57) | -0.04(0.65) | 1.08(0.56) | 1.05(0.53) | 0.61(0.56) | 0.02(0.64) |
| ADNI-EF | 0.71 (0.7) | 0.61 (0.71) | 0.39 (0.70) | 0.03(0.83) | 0.75(0.73) | 0.65(0.69) | 0.39(0.67) | 0.18(0.80) |
| ADNI-LAN | 0.89 (0.7) | 0.76 (0.67) | 0.49(0.71) | 0.21(0.73) | 0.98(0.72) | 0.74(0.68) | 0.50(0.66) | 0.29(0.70) |
| ADNI-VS | 0.24 (0.6) | 0.18 (0.59) | 0.10(0.68) | -0.13 (0.76) | 0.29(0.58) | 0.21(0.58) | 0.07(0.69) | -0.09 (0.79) |
| Plasma Aβ42/40 | - | - | - | - | 0.12(0.01) | 0.12(0.01) | 0.12(0.01) | 0.12 (0.01) |
| Plasma p-tau181 | 15.7 (11.0) | 16.7(16.5) | 16.4(10.2) | 20.2(13.4) | 15.2(11.0) | 15.3(6.7) | 16.1(10.1) | 18.7(9.1) |
| Plasma NFL | 35.6(23.9) | 31.9 (12.6) | 36.6(18.0) | 39.0 (18.8) | 34.8 (14.9) | 32.0(11.8) | 34.6(15.9) | 36.3(15.8) |

### 2.2 Demographics, APOE4, composite cognitive scores

The ADNI baseline demographics (age, gender, years of education), APOE4, and baseline diagnosis were obtained from ADNIMERGE.csv table and the Composite cognitive scores were obtained from “UWNPSYCHSUM.csv”, downloaded from the ADNI website (http://adni.loni.usc.edu/).

.The ADNI study has developed several composite cognitive scores to assess various cognitive domains, including memory, executive function, language, and visuospatial skills. The composite memory score evaluates memory performance using data from tests like the Rey Auditory Verbal Learning Test (RAVLT), AD Assessment Schedule-Cognition (ADAS-Cog), Mini-Mental State Examination (MMSE), and Logical Memory tests [24]. The composite executive function score measures executive function through tests such as the WAIS-R Digit Symbol Substitution, Trails A and B, Digit Span Backwards, Category Fluency, and Clock Drawing tasks [25]. The composite language score assesses language abilities using a variety of tests, including Category Fluency (Animals and Vegetables), Boston Naming, and language tasks from the MMSE, ADAS-Cog, and Montreal Cognitive Assessment (MoCA). The composite visuospatial score evaluates visuospatial skills using clock copying, Constructional Praxis from the ADAS-Cog, and Copy Design from the MMSE [26]. Since these measures were derived by the ADNI study, we refer to them as ADNI-MEM (memory), ADNI-EF (executive function), ADNI-LAN (language), and ADNI-VS (visuospatial functioning) throughout this paper. These composite scores provide standardized, robust measures across cognitive domains, aiding in the early detection and monitoring of cognitive decline in dementia.

To calculate the rate of cognitive decline, we measured the change in each cognitive domain and divided it by the time over which the change occurred. This time frame varied for each individual, as we used the last available follow-up as the endpoint. Additionally, we ensured that the final follow-up was no less than 2 years for all participants.

### 2.3 Plasma biomarkers

In the ADNI study, plasma was collected according to the ADNI procedures manual. Detailed information about the acquisition and analysis methods can be found on the ADNI website available at: adni.loni.usc.edu/methods/documents/.

Plasma A*β*42/40 levels were analyzed by liquid chromatography-tandem mass spectrometry (LC-MS/MS) as previously described [27] and we obtained them from the ADNI depository (files: batemanlab 20190621.csv, batemanlab 20221118.csv). The samples with QC Status “Failed” were excluded. Plasma p-tau181 levels were measured using the Single Molecule Array (Simoa) technique [28] and obtained from the ADNI depository (file: UGOTp-tau181 06 18 20.csv). Plasma NFL was analyzed by the Single Molecule array (Simoa) technique by using a combination of monoclonal antibodies and purified bovine NFL as a calibrator. Plasma NFL was obtained from the ADNI BLENNOWPLASMANFLLONG 10 03 18.csv file.

### 2.4 MRI Measures

As MRI biomarkers, we used the volumetric and thickness measures derived from CAT12 toolbox (https://neuro-jena.github.io/cat/, Version 8.1) using the default settings [29]: For 136 volume measures, the T1-weighted MRI were segmented into grey matter (GM) and white matter (WM) and non-linearly normalized to a stereotactic space using the shooting approach [30]. Based on spatially normalized GM and WM segments, the neuromorphometrics atlas was used to extract the regional volumes. The atlas is derived from the maximum probability tissue labels derived from the “MICCAI 2012 Grand Challenge and Workshop on Multi-Atlas Labeling” http://www.neuromorphometrics.com/2012 MICCAI Challenge Data.htm with the MRIs from OASIS project and the labeled data provided by Neuromorphometrics, Inc. (Neuromorphometrics.com) under academic subscription. The cortical thickness values [31] were computed and registered to the FSaverage surface template [32], where DK40 atlas [33] was used to define 69 regionally averaged cortical thickness measurements.

### 2.5 Machine learning framework

We developed a machine learning framework to predict the rate of cognitive decline based on four composite cognitive scores: memory (ADNI-MEM), executive function (ADNI-EF), language (ADNI-LAN), and visuospatial (ADNI-VS) abilities. The framework involves two main stages. In the first stage, ridge linear regression (RLR) was applied to MRI data to calculate an MRI score. In the second stage, the MRI score is combined with other predictors including demographics, APOE4, baseline composite cognitive scores, and plasma measures, using random forest regression. To ensure robust model performance, we employed nested and stratified cross-validation with two loops. The inner CV loop was used for calculating MRI score and the outer CV loop was used to split data into main training and test sets. This approach allowed us to build the MRI model while ensuring that the same data was not used for both training and predicting MRI scores in the training set, thus preventing overfitting. This two-stage process is illustrated in Fig. 1. The same framework was used to predict the rate of cognitive decline based on the four composite cognitive scores. The participants in all experiments were the same, and the experiments were conducted in parallel for Cohort 1 and Cohort 2.

**Fig. 1.**
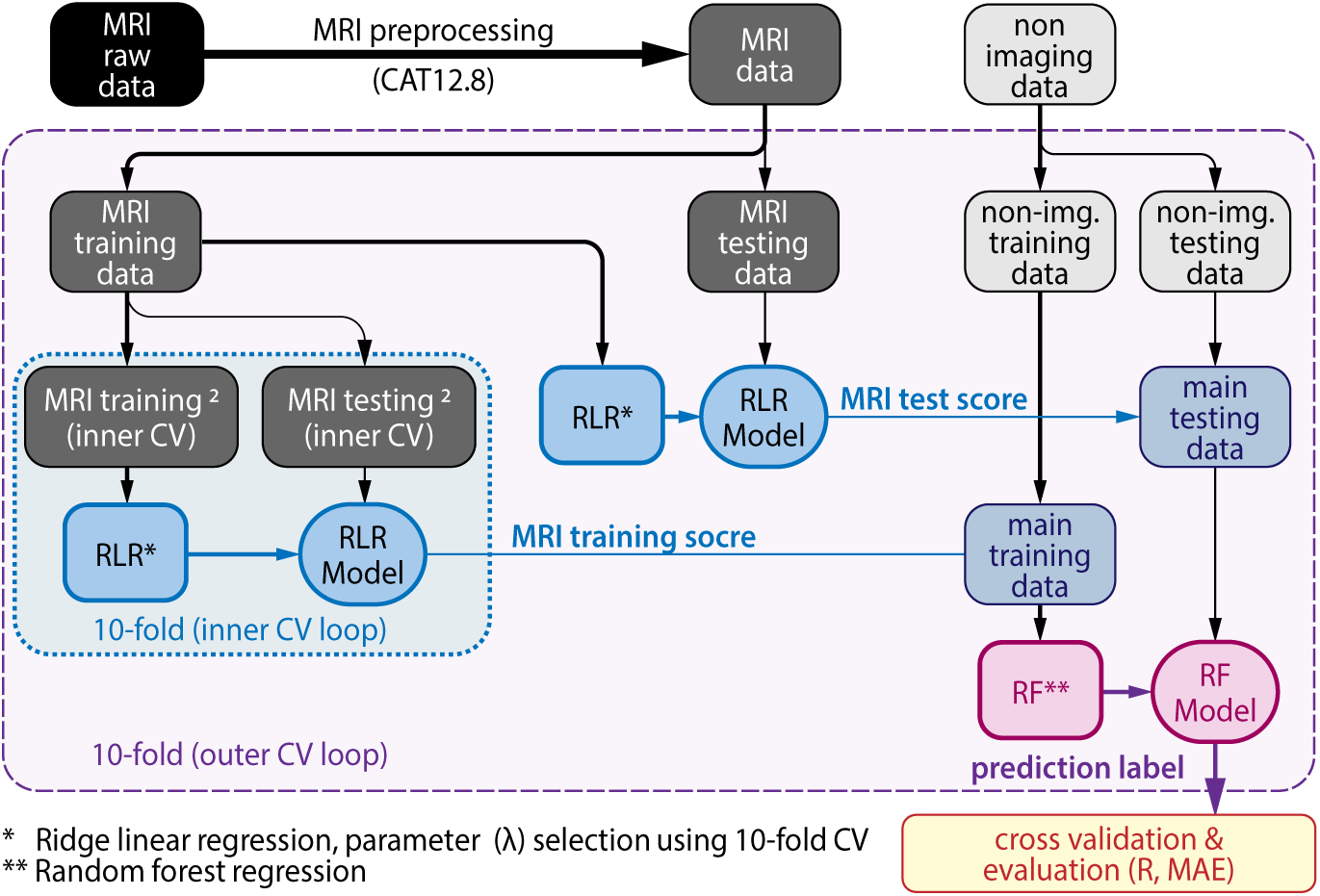
Schematic representation of the regression framework.

We designed four models with different feature combinations. The first model, the basic model, was trained using only demographic information, APOE4, and the baseline composite cognitive scores. The second model, the plasma model, included plasma biomarkers and predictors used in the basic model. The third model, the MRI model, incorporated MRI data with the predictors of the basic model. Finally, the fourth model, termed the combined model, utilized all available predictors: demographic data, APOE4, composite cognitive scores, plasma biomarkers, and MRI data. Our two-stage framework, illustrated in Fig. 1, was applied to experiments involving MRI measures (i.e., the MRI and combined models). For the basic and plasma models, which did not include MRI data, we used a simple random forest regression to predict the rate of cognitive decline. The R codes are available at “https://github.com/ElahehMoradi/Cognitive-decline-prediction”.

### 2.6 Implementation and performance evaluation

To divide the data into training and test sets, we used two nested and stratified cross-validation loops, each with 10 folds. In the inner CV loop, the MRI-training set (outer loop) was further divided into MRI-training and test sets (inner CV) to learn the MRI model with the RLR approach and predict MRI scores for the training set (outer loop). We used the inner CV loop to ensure that the same dataset was not used for both learning the MRI model and calculating the MRI score in the training set, thus avoiding overfitting. After calculating the MRI scores for the training data (outer loop), the MRI training set of the outer CV loop was used to predict the MRI score for the MRI test set of the outer CV loop. The MRI scores of the training data were then combined with other predictors of the training dataset, and the MRI score of the test set was combined with other predictors of the test set. We then used the new training set with all the predictors, including the MRI score, to design a Random Forest regression model and predict the rate of cognitive decline in the test set. The Random Forest regression parameters were set to their default values, except for the number of trees, which was increased to 1,000 to enhance the model’s stability and improve the accuracy of feature importance determination. It is important to note that the test set of the outer loop was not used in any learning stages and was used only for evaluating the model. The implementation steps are visualized in Fig. 1.

The performance of the model was evaluated based on the cross-validated Pearson correlation coefficient (R) and the mean absolute error (MAE) between the estimated and true rate of cognitive decline. The reported results are averages over 10 nested 10-fold CV runs to minimize the effect of random variation.

To compare the correlation coefficient we used methods described by Diedenhofen and colleagues [34]. All the analyses were done using R (version 4.1.1), with the following packages: glmnet [35], caret [36], cocor [37], pROC [38], Daim [39], ggplot2 [40], and complexheatmap [41], survival [42].

## 3 Results

### 3.1 Predicting the rates of cognitive decline in non-demented individuals

We predicted the rate of cognitive decline in different cognitive domains based on ADNI-MEM, ADNI-EF, ADNI-LAN, and ADNI-VS composite cognitive scores. As explained in the Methods, we designed four models with different feature combinations for each regression experiment. Fig. 2 shows the results of all these computational analyses. These results are the average over 10 repeated 10-fold cross-validation analyses for each experiment.

**Fig. 2.**
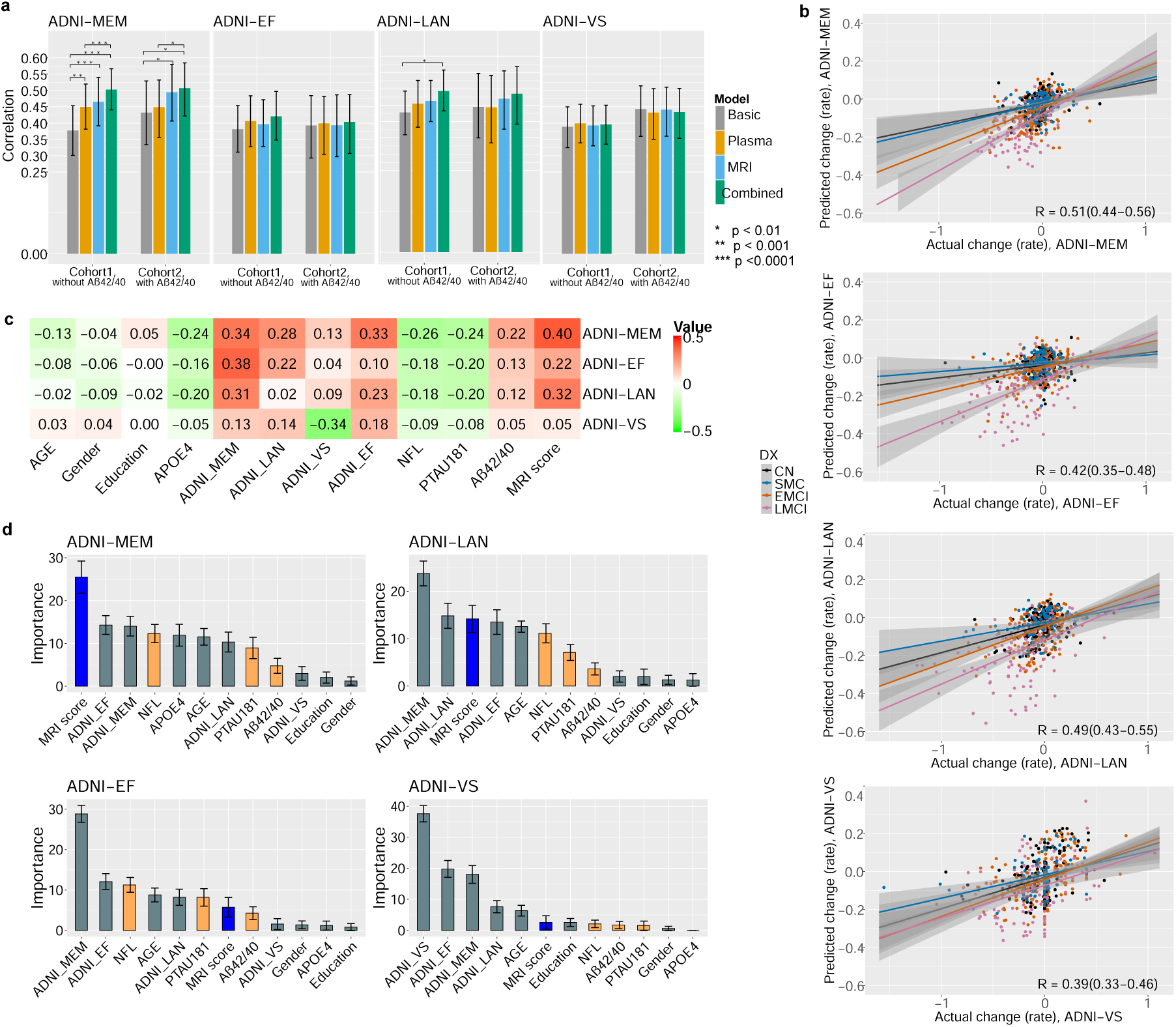
Predicting the rates of cognitive decline in non-demented individuals: a) Bar plots with 95% confidence interval error bars show the average correlation score across 10 computation runs for predicting the rate of cognitive decline in both study cohorts. b) Scatter plot for actual cognitive decline rate vs. predicted rate using the combined model in Cohort 1. c) Heatmap of correlation values derived from the univariate analysis of the actual rate of cognitive decline and the predictors based on all four composite cognitive scores in Cohort 2. d) The importance of different predictors calculated by RF regression models for each experiment using the combined model in Cohort 2, which includes all predictors, including plasma A*β*42/40. (MRI score is the mean value of cross-validated MRI score derived by applying RLR on MRI data in test subset).

According to Fig. 2 (panel a), the prediction performance in cohorts 1 and 2 was similar, despite the absence of A*β*42/40 in Cohort 1. The following analysis is based on Cohort 1 due to the larger sample size. For predicting the rate of change in the ADNI-MEM measure, the average correlation score across 10 computation runs, derived from the basic model, was 0.38 with a 95% confidence interval (CI) of 0.30 to 0.45. The average MAE was 0.11 (95% CI of 0.10 to 0.12). Adding plasma biomarkers to the basic model (plasma model) significantly improved performance, with an improved correlation score of 0.45 (95% CI of 0.38 to 0.52) and a decreased MAE of 0.10 (95% CI of 0.10 to 0.11). Similarly, adding MRI data to the basic model (MRI model) significantly improved performance to a correlation score of 0.46 (95% CI of 0.39 to 0.54) and an MAE of 0.10 (95% CI of 0.10 to 0.11). Although the correlation score of the MRI model was slightly higher than that of the plasma model, the difference was not statistically significant (p = 0.43). Finally, the combined model integrating plasma and MRI measures to the basic model, provided significantly improved prediction performance compared to all other models (basic, plasma, and MRI models), with an average correlation score of 0.50 (95% CI of 0.44 to 0.57) and an MAE of 0.10 (95% CI of 0.09 to 0.11). The improvement in the correlation score of the combined model compared to the plasma and MRI models indicates that plasma and MRI measures provide different information for predicting the rate of cognitive decline based on the ADNI-MEM score, and both data types are important for this prediction task.

Interestingly, within other cognitive domains (ADNI-EF, ADNI-LAN, ADNI-VS), the cognitive changes could be predicted as well based on the basic model than based on the model integrating biomarker information (see Fig. 2 a). The only exception to this was ADNI-LAN where the combined model offered a significant advantage over the basic model.

Fig. 2 panel b presents a scatter plot of the actual cognitive decline rate versus the predicted rate using the combined model in Cohort 1. Individuals from different diagnostic groups are labeled with different colors, and a linear fit is plotted for each diagnosis group. In the ADNI-MEM score, a clear relationship between the rate of cognitive decline and the diagnostic groups existed. As expected, there was a higher correlation between the observed and predicted values in the MCI groups (EMCI and LMCI) compared to cognitively unimpaired (CU) individuals (CN and SMC). It is also evident that the rate of cognitive change was markedly smaller in the CU group, catalyzing more challenging prediction task in these individuals. This effect is also visible in the ADNI-EF and, on a smaller scale, in the ADNI-LAN scores, but not in the ADNI-VS score.

We investigated the contribution of individual variables in predicting the rate of cognitive decline. Fig. 2, panel d, shows the importance of different variables derived from 100 RF models, based on 10 runs of 10-fold cross-validation. The bar plot displays the mean importance of each variable across 100 computation runs. These results are from the combined model using Cohort 2, which includes the A*β*42/40 measure. The MRI score represents the mean value of the cross-validated MRI score, derived from RLR applied to the MRI data in the test subset, as described in the methods section.

For predicting the rate of cognitive decline based on the ADNI-MEM score, the MRI score emerged as the most important predictor. While it did not hold the highest importance in other cognitive domains, it still played a significant role in predicting ADNI-LAN performance and, to a lesser extent, in the ADNI-EF score. Interestingly, APOE4 was found to be significant only for ADNI-MEM, with no notable importance for other cognitive scores. Furthermore, the baseline ADNI-MEM score was the strongest predictor for cognitive decline rate in the ADNI-EF and ADNI-LAN domains, surpassing the predictive value of the baseline ADNI-EF and ADNI-LAN scores.

Among the plasma biomarkers, A*β*42/40 had the lowest importance in predicting the rate of cognitive decline across different composite cognitive scores. This might explain why the prediction performance in Cohort 1 and Cohort 2 was similar, despite the absence of the A*β*42/40 measure in Cohort 1. In contrast, plasma NFL and p-tau181 were identified as important features for predicting the rate of cognitive decline based on ADNI-MEM, ADNI-EF, and ADNI-LAN scores, but not for ADNI-VS score. In the case of ADNI-VS, all plasma biomarkers and MRI scores had low importance, indicating that baseline composite cognitive scores contributed the most to the model. Univariate analysis of the actual rate of cognitive decline with different predictors revealed that neither plasma biomarkers nor MRI scores showed significant correlations with ADNI-VS (Fig. 2, panel c). This finding aligns with the low contribution of these variables to the ADNI-VS model. In contrast, plasma biomarkers and MRI measures demonstrated stronger correlations with ADNI-MEM and, to a certain degree, with ADNI-EF and ADNI-LAN, which explains their greater contribution to model performance in these cognitive domains.

### 3.2 Predicting the rates of cognitive decline in cognitively unimpaired and MCI groups separately

We extended our analysis by predicting the rate of cognitive decline in CU and MCI groups separately. Participants in both cohorts were divided into cognitively unimpaired (CN and SMC) and MCI groups (EMCI and LMCI), and the combined method was applied to each group individually. The results are illustrated in Fig. 3. As expected, predicting the rate of cognitive decline was markedly more challenging in the CU individuals compared to the MCI individuals across all four cognitive domains. This difference is particularly substantial and significant in ADNI-MEM and ADNI-EF scores.

**Fig. 3.**
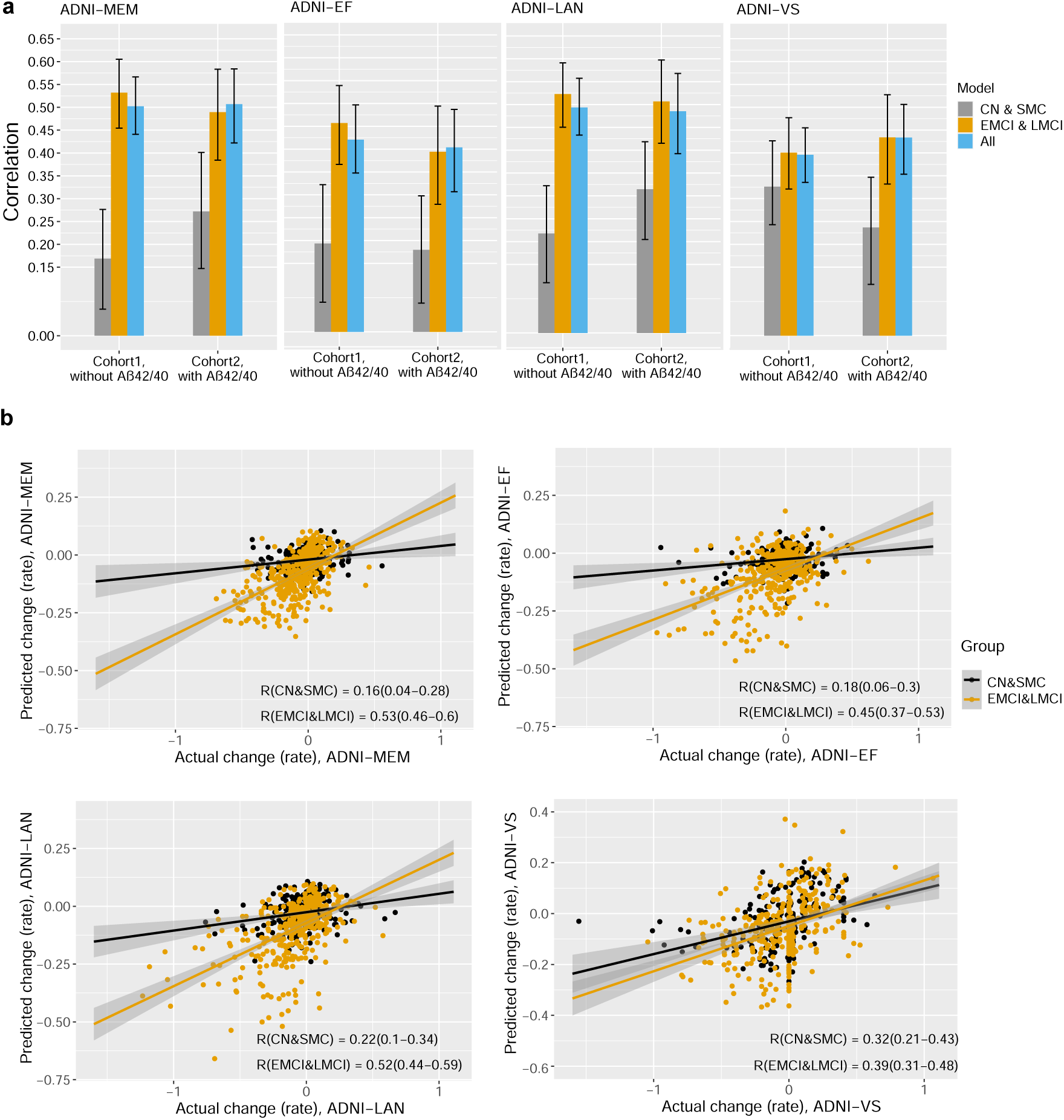
Predicting the rates of cognitive decline in CU and MCI groups separately. a) Bar plots with 95% confidence interval error bars show the average correlation score across 10 computation runs for predicting the rate of cognitive decline in CU and MCI groups, both separately and combined, in Cohort 1. b) Scatter plot for actual cognitive decline rate vs. predicted rate using the combined model in Cohort 1 in CU and MCI groups.

For predicting the rate of cognitive decline in the ADNI-MEM domain, the average correlation score across 10 computation runs in Cohort 1 was 0.17 (95% CI of 0.06 to 0.28) for the CU group and 0.53 (95% CI of 0.45 to 0.61) for the MCI group. For ADNI-EF, the correlation score was 0.19 (95% CI of 0.07 to 0.32) for the CU group and 0.46 (95% CI of 0.37 to 0.54) for the MCI group. In the case of ADNI-LAN, the differences in correlation between predicted and actual rates in the CU and MCI groups were slightly smaller but still significant. The correlation score based on ADNI-LAN was 0.22 (95% CI of 0.11 to 0.32) for the CU group and 0.52 (95% CI of 0.45 to 0.59) for the MCI group. The results for ADNI-VS differed from other cognitive scores, with a significantly smaller difference in correlation scores in CU and MCI groups that was not statistically significant based on confidence intervals. For ADNI-VS, the correlation score was 0.33 (95% CI of 0.24 to 0.42) for the CU group and 0.40 (95% CI of 0.32 to 0.47) for the MCI group.

Fig. 3b shows the scatter plot for one computation run with median performance across 10 computation runs for actual versus predicted rates of cognitive decline based on different composite cognitive scores, with a linear fit plotted for each group. A clear difference can be seen for ADNI-MEM, ADNI-EF, and ADNI-LAN scores between the CU and MCI groups, but not for the ADNI-VS score.

### 3.3 Prediction of progression to MCI/dementia

To assess the effectiveness of our cognitive decline prediction models in predicting dementia progression, we evaluated the predictive power of estimated cognitive decline rates, derived from various composite cognitive scores. This analysis was conducted across both CU individuals and those with MCI.

In Cohort 2, which included 160 cognitively unimpaired individuals (CN and SMC), 30 transitioned to MCI or dementia during the follow-up period, and 96 remained stable with a follow-up duration of at least four years. In the MCI group (EMCI and LMCI), 65 individuals progressed to dementia, and 120 remained with MCI status.

For the analysis, predicted cognitive decline rates were categorized into 3 tertiles. Individuals in the first tertile had the lowest predicted rate of cognitive decline, while those in the last tertile had the highest predicted rate. Fig. 4 shows the Kaplan-Meier survival analysis results and log-rank tests across tertiles, focusing on the predicted rate of the ADNI-MEM cognitive score for Cohort 2. Results for other cognitive scores and experiments with Cohort 1 are provided in the supplementary materials (Supplementary Fig. 2-5). The survival curves reveal significant differences among the tertiles in both CU and MCI individuals, as determined using all four models (log-rank P *<* 0.001).

**Fig. 4.**
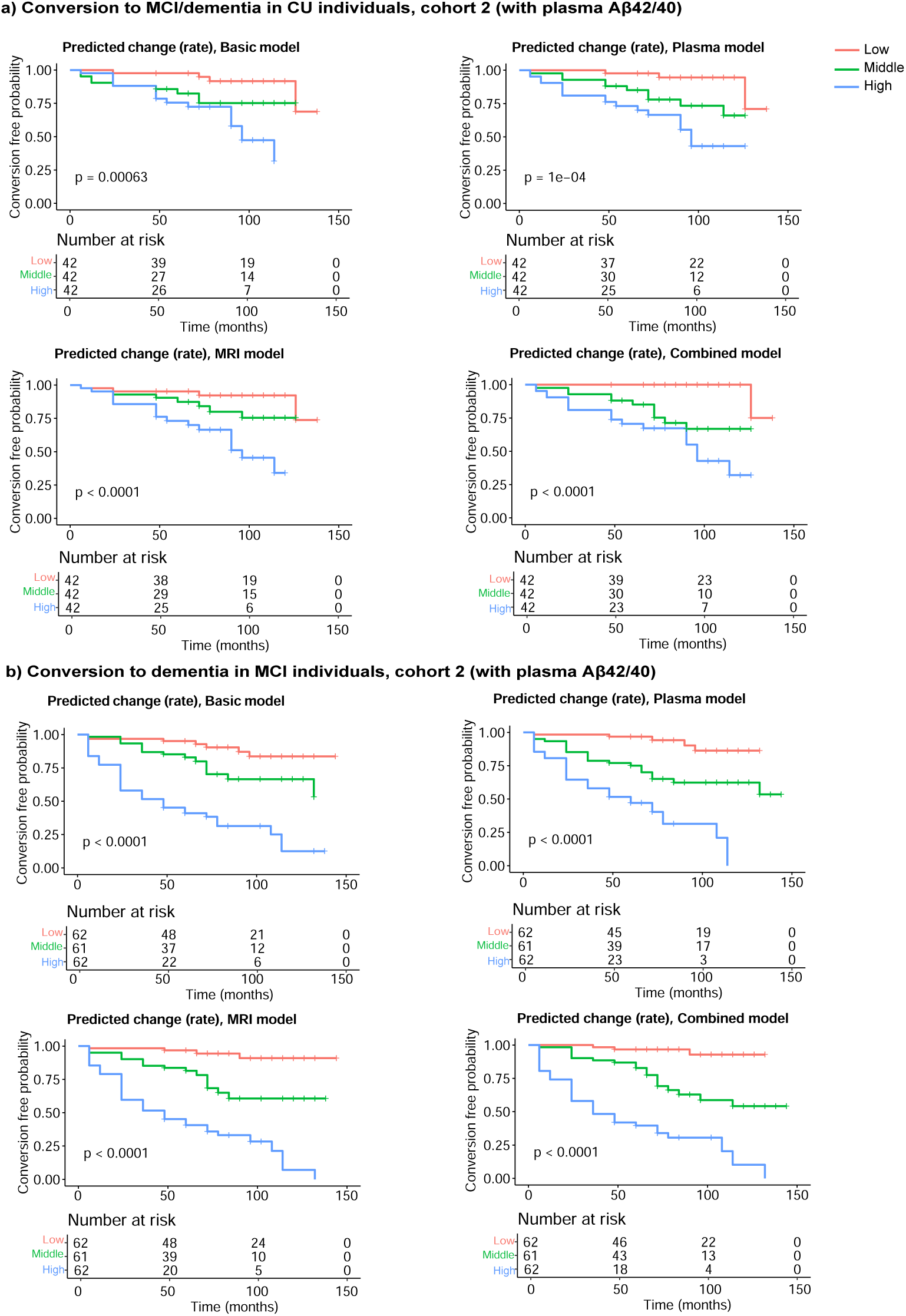
Kaplan-Meier survival curves for conversion to MCI/AD in CU and MCI groups in cohort 2: The predicted rate of cognitive decline in ADNI-MEM score was divided into 3 tertiles (low, middle, high). Vertical tick marks on lines indicate times at which the individual was censored. p-values are for log-rank tests among the tertiles. a) in CU individuals for converting to MCI/dementia, b) in MCI individuals for converting to dementia. Basic model: Demographics, APOE4, Composite cognitive scores. Plasma model: Demographics, APOE4, Composite cognitive scores, plasma biomarkers. MRI model: Demographics, APOE4, Composite cognitive scores, MRI data. Combined model: Demographics, APOE4, Composite cognitive scores, plasma biomarkers, MRI data.

The predicted ADNI-MEM score rate from the basic model demonstrated a C-index (concordance index) of 0.68 for predicting progression to MCI/dementia in CU individuals. When plasma biomarkers were added (plasma model), the C-index improved to 0.72. The MRI model showed a slightly lower C-index of 0.69. However, the combined model yielded an improvement over the plasma model, achieving a C-index of 0.76, indicating better predictive accuracy. For predicting dementia progression in individuals with MCI, the basic model yielded a C-index of 0.76. Adding plasma biomarkers did not enhance predictive accuracy (C-index of 0.74), but the MRI model offered a slight improvement with a C-index of 0.77. The combined model also achieved the highest performance for the MCI group, with a C-index of 0.79. Although our cognitive decline prediction model based on ADNI-MEM was primarily designed to estimate memory domain decline rates, rather than directly predict dementia conversion, it still showed strong predictive power for early-stage dementia, even in CU individuals.

Table 2 summarizes the hazard ratios (HRs) for the predicted rates of cognitive decline, as measured by the ADNI-MEM score, across tertiles using four models: basic, plasma, MRI, and combined. The first tertile, representing individuals with the lowest predicted rate of decline, served as the reference group. Among CU individuals, the combined model demonstrated that those in the highest tertile (fastest predicted decline) had a risk of progressing to MCI/dementia that was more than 33 times greater than those in the lowest tertile (slowest predicted decline). For the MRI and plasma models, the risks were 7.9 and 9.7 times higher, respectively. Similarly, for MCI individuals, the combined model showed that those in the highest tertile had a risk of progressing to dementia that was over 29 times greater compared to the lowest tertile. The MRI and plasma models indicated risks of 21 and 14 times higher, respectively. The results in Cohort 1 were also similar to those in Cohort 2 (Supplementary Fig. 2).

**Table 2.**
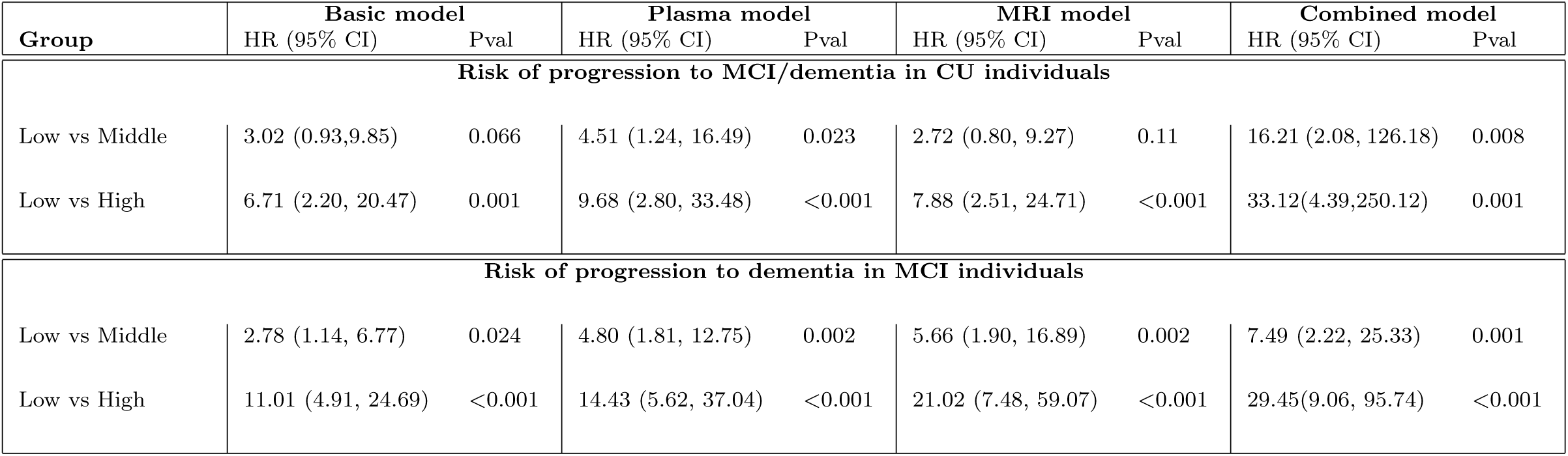
Progression risk to dementia. Hazard ratios (HR) (95% CI) were calculated using Cox regression analysis. The first quartile is used as a reference for calculating HR.

The predicted rate of cognitive decline in other cognitive domains was not as strong as the one based on ADNI-MEM for predicting progression to dementia, especially in the early stages. The survival curves for the predicted rate of cognitive declines based on ADNI-EF, ADNI-VS, and ADNI-LAN show significant differences among the three groups in MCI individuals, as determined using all four models (log-rank P *<* 0.001), but not in CU individuals (log-rank P *<* 0.001) (Supplementary Fig. 3-5).

## 4 Discussion

The objective of this study was twofold: first, to investigate the role of plasma biomarkers, in combination with other non-invasive measures, in predicting the rate of cognitive decline in individuals without dementia; and second, to predict the rate of decline across different cognitive domains—memory, executive function, language, and visuospatial abilities—and assess their associations with progression to dementia. To achieve this, we developed a two-stage machine learning framework. In the first stage, regularized logistic regression (RLR) was applied to MRI data to derive an MRI score. In the second stage, the MRI score was combined with plasma biomarkers, demographics, APOE4, and baseline composite cognitive scores using random forest regression.

Our findings indicate that plasma biomarkers are effective in predicting distinct patterns of domain-specific cognitive decline and can potentially be used to predict progression to dementia, even in cognitively unimpaired individuals. While plasma biomarkers may not fully capture all the pathological changes occurring in the brain, they serve as a non-invasive source of information, offering valuable insights into disease mechanisms before the clinical onset of dementia [8, 10, 11, 13, 19, 21].

We conducted multiple experiments with various feature combinations to identify the influential biomarkers in predicting the rate of cognitive decline. Notably, our findings indicated that the ADNI-MEM-based predictive model significantly benefited from the inclusion of additional features, while the performance of other composite cognitive measures showed little improvement with these additions. Specifically, for predicting the rate of cognitive decline based on the ADNI-MEM score, the correlation between the actual and predicted rates improved significantly (P *<* 0.001), with the correlation score increasing from 0.38 in the basic model to 0.45 in the plasma model after adding plasma biomarkers to demographics, APOE4, and baseline composite cognitive scores. Additionally, incorporating MRI data into the plasma model to form the combined model further improved the prediction performance, with the correlation score rising significantly (P *<* 0.001) to 0.50. However, for other composite cognitive scores, adding plasma or MRI measures to the basic model did not result in statistically significant improvements (P *>* 0.01). The exception was the ADNI-LAN score, where the combined model showed a significant improvement (correlation score increased to 0.49, P = 0.005) over the basic model (correlation score 0.43), although this improvement was not statistically significant in the MRI model (correlation score 0.46) or the plasma model (correlation score 0.45).

Assessing the importance of different features in the combined model revealed that while plasma biomarkers were useful for predicting the rate of cognitive decline, they were not the top predictors. Instead, the MRI and baseline composite cognitive scores were the strongest predictors, along with plasma biomarkers NfL and p-tau 181. Among the plasma biomarkers, the A*β*42/40 ratio was less predictive than NfL and p-tau 181. Notably, NfL consistently ranked among the top four predictors for memory (ADNI-MEM) and executive function (ADNI-EF) decline. This aligns with recent studies highlighting the limited predictive value of amyloid-beta measures compared to markers of neuronal injury and tau pathology. Both NfL and p-tau 181 are linked to faster cognitive decline and increased risk of progression to dementia [20, 21]. Interestingly, for the visuospatial domain (ADNI-VS), baseline cognitive scores were the most important predictors, while MRI, plasma biomarkers, and APOE4 had minimal impact. These findings highlight that the predictive value of biomarkers varies across cognitive domains.

We further investigated the predictive performance of cognitive decline rates across different domains separately for CU and MCI groups. By analyzing datasets with varying levels of memory impairment, we could determine the relationship between each cognitive measure and dementia. The results showed strong associations between the information detected by ADNI-MEM, ADNI-EF, and ADNI-LAN scores for different diagnostic groups, but not ADNI-VS. Prediction performance was significantly worse in the CU group than the MCI group, which aligns with our expectations given the lower rate of cognitive decline in CU individuals, making prediction more challenging. No significant association between ADNI-VS and diagnostic groups was observed.

We also assessed the effectiveness of our prediction models for cognitive decline across different domains in predicting dementia progression in CU and MCI groups using survival analysis. Across all models, which varied in feature combinations and composite cognitive scores, significant predictive value was demonstrated for dementia progression in the MCI group. Notably, the predicted rate based on the ADNI-MEM score demonstrated the highest predictive power, while those based on the ADNI-VS score showed the lowest. In contrast, predicting progression to MCI/dementia in CU individuals proved more challenging. Only the predicted rate based on ADNI-MEM showed significant predictive power for dementia progression in this group. Among the models tested, the combined model demonstrated the highest predictive accuracy. Since memory impairment is often the earliest symptom of AD, driven by neurodegeneration in regions such as the hippocampus, the ADNI-MEM-based predictions were particularly effective in identifying progression to MCI or dementia, even in individuals without cognitive impairment.

Previous research on plasma biomarkers in AD has primarily focused on detecting brain amyloidosis, often using the plasma A*β*42/40 ratio to detect PET or CSF A*β* positivity [7, 10, 19]. Fewer studies, however, have explored the role of plasma biomarkers in predicting cognitive decline. For example, Mattsson et al.[8] evaluated combinations of plasma biomarkers, including p-tau 217, to predict cognitive decline in A*β*-positive CU individuals using MMSE and modified Preclinical Alzheimer Cognitive Composite (mPACC) scores. Their findings highlighted the strong predictive power of plasma p-tau 217 in detecting cognitive decline in preclinical AD patients. Similarly, Zhang et al. [11] investigated the association of plasma biomarkers such as A*β*42/40, p-tau 181, NfL, and glial fibrillary acidic protein (GFAP) with cognitive decline across different domains. Their study showed that memory decline was most strongly associated with p-tau 181, while attention, executive function, and visuospatial abilities were more closely linked to NfL levels. A*β*42/40 emerged as the most efficient marker for distinguishing memory decline, whereas GFAP was particularly effective in identifying decline patterns in language and visuospatial functions. Our study shares similarities with Zhang et al. [11] in assessing cognitive decline across domains but differs in methodology. We employed machine learning for predictive modeling, prioritizing high-accuracy forecasting of future outcomes, rather than exploratory analysis aimed at understanding relationships and generating hypotheses [43]. Machine learning allows for identifying complex data patterns that conventional methods may overlook. Additionally, we assessed the impact of combining plasma biomarkers with other non-invasive measures to improve predictive accuracy. Finally, we evaluated the effectiveness of our predictive models in forecasting conversion to MCI or dementia in CU and MCI groups. These analyses offer a more nuanced understanding of the potential of plasma biomarkers in early AD detection.

Our findings emphasize the strengths and limitations of plasma biomarkers in dementia research. While they excel in predicting memory decline and dementia progression, their predictive power is limited in non-memory domains. This may be due to the focus on AD-specific plasma biomarkers, i.e., A*β*42/40, p-tau 181, and NfL, which primarily reflect amyloid plaques, tau aggregation, and neurodegeneration—core pathological features of AD. These biomarkers are highly relevant for detecting AD-related pathology and demonstrate stronger predictive value for cognitive decline in memory-related domains. However, they tend to be less predictive in non-memory domains like visuospatial abilities, where neurodegeneration may not be as directly influenced by amyloid or tau pathology. This likely explains the reduced effectiveness of plasma biomarkers in predicting cognitive decline in these areas and their limited performance in early AD prediction.

In conclusion, our study demonstrates the potential of plasma biomarkers, especially when combined with non-invasive measures such as MRI data and cognitive scores, to predict domain-specific cognitive decline and progression to dementia. Our findings also highlight the challenge of predicting cognitive decline in the early stages of AD, particularly in individuals who are still cognitively unimpaired. However, when CU individuals are analyzed alongside those who have slightly progressed toward AD, such as those in the MCI stage, ML algorithms can help predict cognitive changes to some extent and even predict progression to MCI/dementia with a certain level of accuracy. This has significant clinical implications, as current methods struggle to accurately predict cognitive decline in CU individuals—a critical area given the development of new AD-modifying drugs that should be administered in the early stages of the disease.

A possible limitation of this study is its focus on AD-specific biomarkers, which restricted our ability to explore relationships between other pathologies and types of dementia across different cognitive domains. Moreover, since our experiments were conducted using the ADNI cohort, which predominantly focuses on AD, the results may not generalize well to non-memory domains where AD-specific biomarkers are less relevant. The memory domain is most closely associated with AD, which explains why our predictions were more accurate in this area. These findings underscore the need for future studies that incorporate a broader range of biomarkers and pathologies to enhance understanding and prediction of cognitive decline across various types of dementia.

## Data Availability

Data used in the preparation of this article
were obtained from the Alzheimer's Disease Neuroimaging Initiative (ADNI)
database (https://adni.loni.usc.edu/). Details about data access are detailed there.
The participant's RIDs are provided in Supplementary Materials

## Declarations

### Ethics approval and consent to participate

The ADNI study was approved by the Institutional Review Board (IRB) of each participating site and was conducted in accordance with Federal Regulations. For participants in ADNI protocols, written informed consent was obtained and the study was conducted in accordance with the Declaration of Helsinki.

### Consent for publication

Not applicable.

### Availability of data and materials

Data used in the preparation of this article were obtained from the Alzheimer’s Disease Neuroimaging Initiative (ADNI) database (https://adni.loni.usc.edu/). Details about data access are detailed there. The participant’s RIDs are provided in Supplementary Materials.

## Acknowledgments

Data collection and sharing for this project was funded by the Alzheimer’s Disease Neuroimaging Initiative (ADNI) (National Institutes of Health Grant U01 AG024904) and DOD ADNI (Department of Defense award number W81XWH-12-2-0012). ADNI is funded by the National Institute on Aging, the National Institute of Biomedical Imaging and Bioengineering, and through generous contributions from the following: AbbVie, Alzheimer’s Association; Alzheimer’s Drug Discovery Foundation; Araclon Biotech; BioClinica, Inc.; Biogen; Bristol-Myers Squibb Company; CereSpir, Inc.; Cogstate; Eisai Inc.; Elan Pharmaceuticals, Inc.; Eli Lilly and Company; EuroImmun; F. Hoffmann-La Roche Ltd and its affiliated company Genentech, Inc.; Fujirebio; GE Healthcare; IXICO Ltd.; Janssen Alzheimer Immunotherapy Research & Development, LLC.; Johnson & Johnson Pharmaceutical Research & Development LLC.; Lumosity; Lundbeck; Merck & Co., Inc.; Meso Scale Diagnostics, LLC.; NeuroRx Research; Neurotrack Technologies; Novartis Pharmaceuticals Corporation; Pfizer Inc.; Piramal Imaging; Servier; Takeda Pharmaceutical Company; and Transition Therapeutics. The Canadian Institutes of Health Research is providing funds to support ADNI clinical sites in Canada. Private sector contributions are facilitated by the Foundation for the National Institutes of Health ( www.fnih.org). The grantee organization is the Northern California Institute for Research and Education, and the study is coordinated by the Alzheimer’s Therapeutic Research Institute at the University of Southern California. ADNI data are disseminated by the Laboratory for Neuro Imaging at the University of Southern California.

During the preparation of this work, the authors used GPT-4 from OpenAI in order to improve language and readability. After using this tool/service, the authors reviewed and edited the content as needed. The authors take full responsibility for the content of the publication.

