## Supplementary material for "Integrating plasma, MRI, and cognitive biomarkers for personalized prediction of decline across cognitive domains": Supplemenatry Materials: Supplementary Materials.pdf

December 31, 2024

### Spplementary Figures

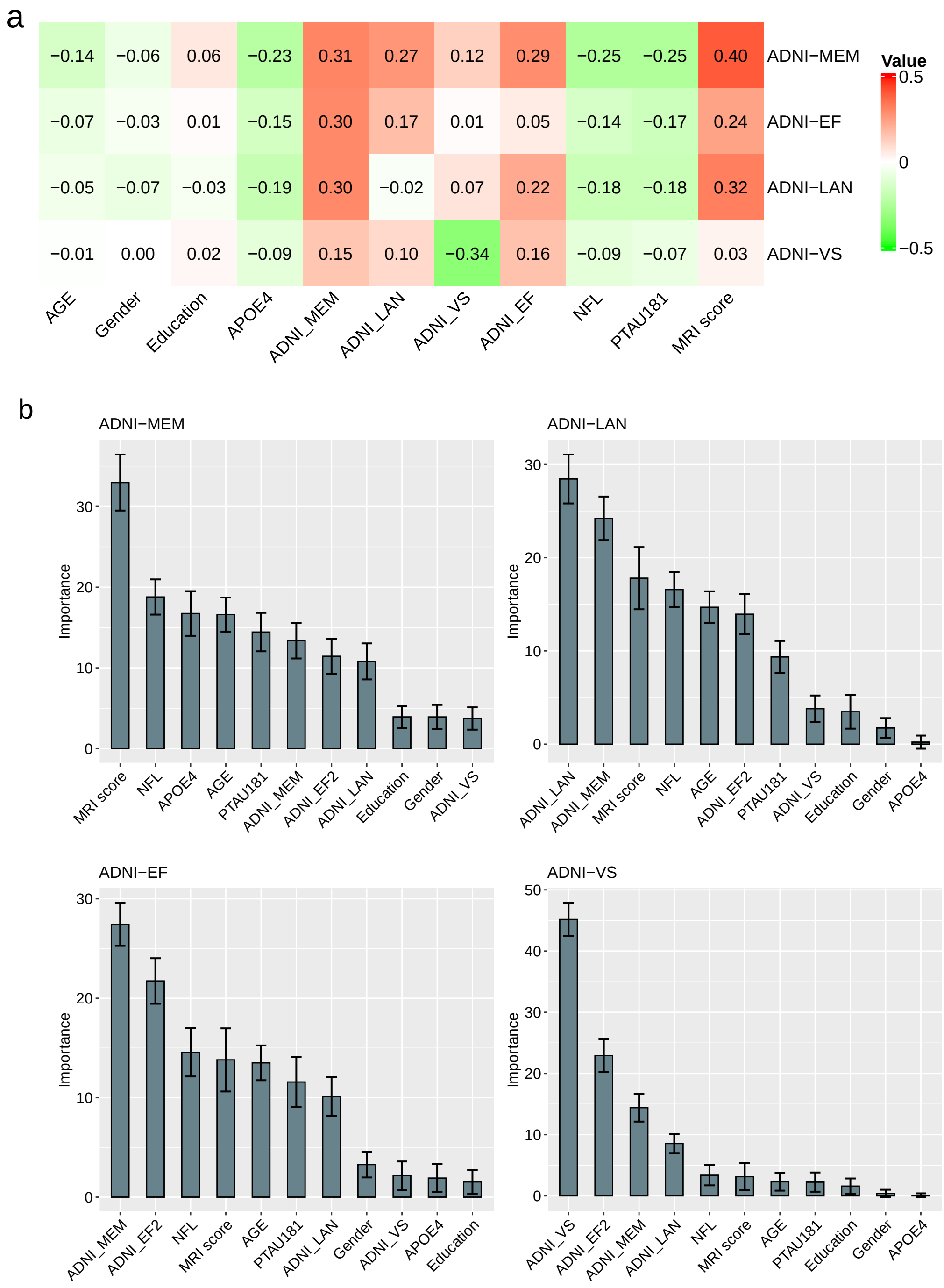


Figure S1: Predicting the rates of cognitive decline in non-demented individuals, results from Cohort 1:a) Heatmap of correlation values derived from the univariate analysis of the actual rate of cognitive decline and the predictors based on all four composite cognitive scores.b) The importance of different predictors calculated by RF regression models for each experiment using the combined model, which includes all predictors, except plasma A$\beta$42/40. (MRI score is the mean value of cross-validated MRI score derived by applying RLR on MRI data in test subset).

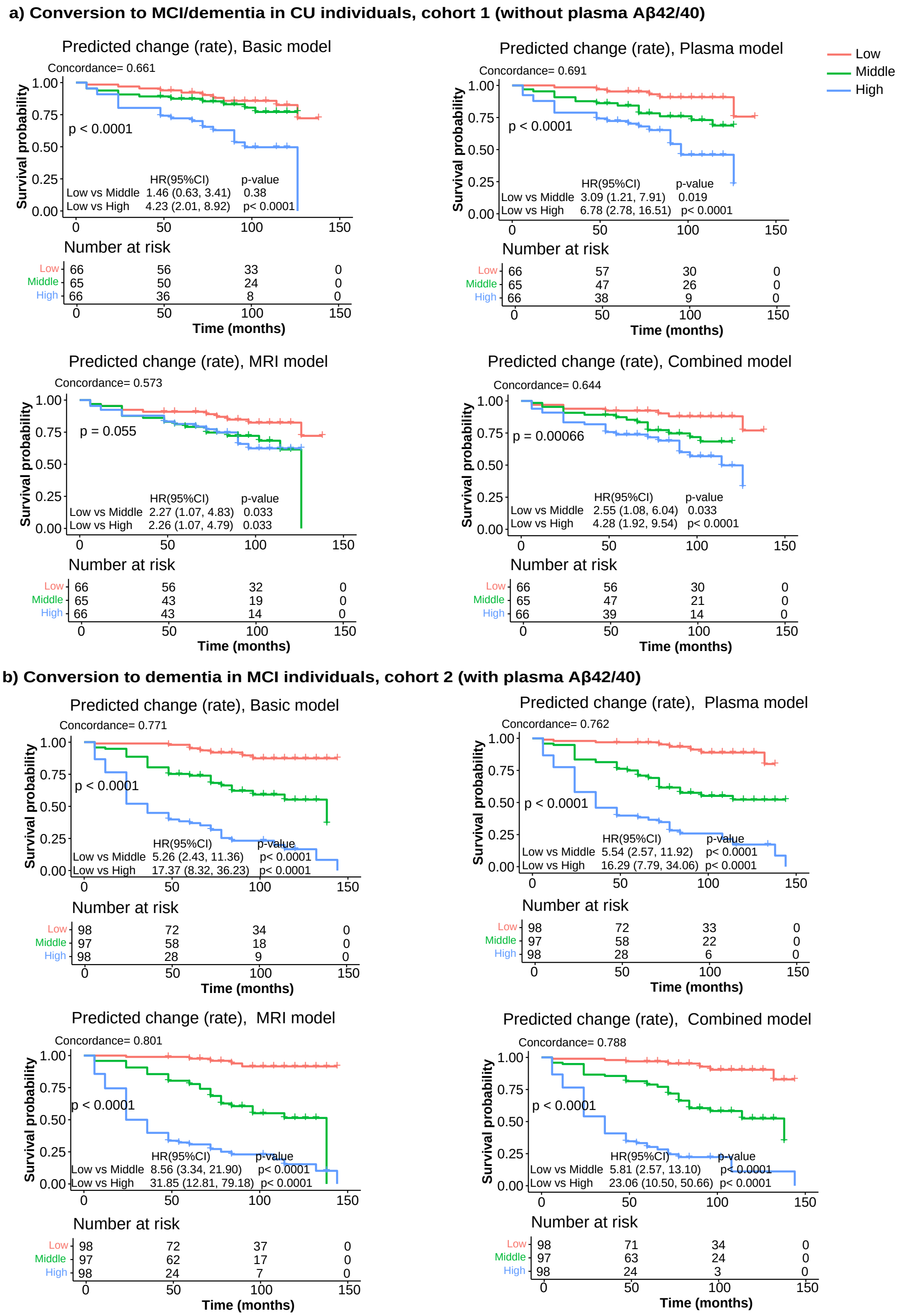


Figure S2: Kaplan-Meier survival curves for conversion to MCI/AD in CU and MCI individuals in cohort 1: The predicted rate of cognitive decline in ADNI-MEM score was divided into 3 tertiles (low, middle, high). Vertical tick marks on lines indicate times at which the individual was censored. p-values are for log-rank tests among the tertiles. Plasma model: Demographics, APOE4, Composite cognitive scores, plasma biomarkers. MRI model: Demographics, APOE4, Composite cognitive scores, MRI data. Combined model: Demographics, APOE4, Composite cognitive scores, plasma biomarkers, MRI data.

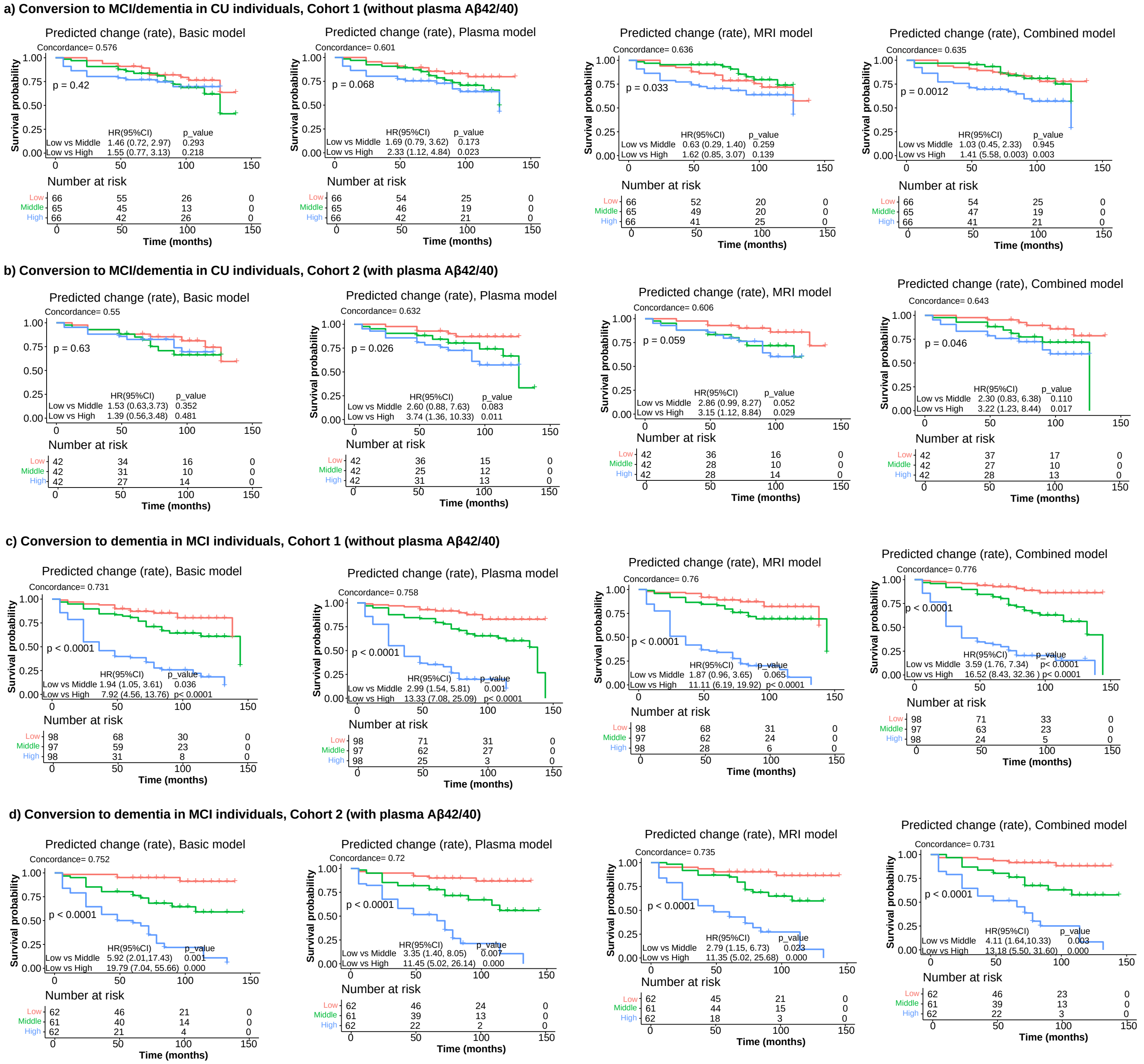


Figure S3: Kaplan-Meier survival curves for conversion to MCI/AD in CU and MCI individuals: The predicted rate of cognitive decline in ADNI-EF score was divided into 3 tertiles (low, middle, high). Vertical tick marks on lines indicate times at which the individual was censored. p-values are for log-rank tests among the tertiles. Plasma model: Demographics, APOE4, Composite cognitive scores, plasma biomarkers. MRI model: Demographics, APOE4, Composite cognitive scores, MRI data. Combined model: Demographics, APOE4, Composite cognitive scores, plasma biomarkers, MRI data.

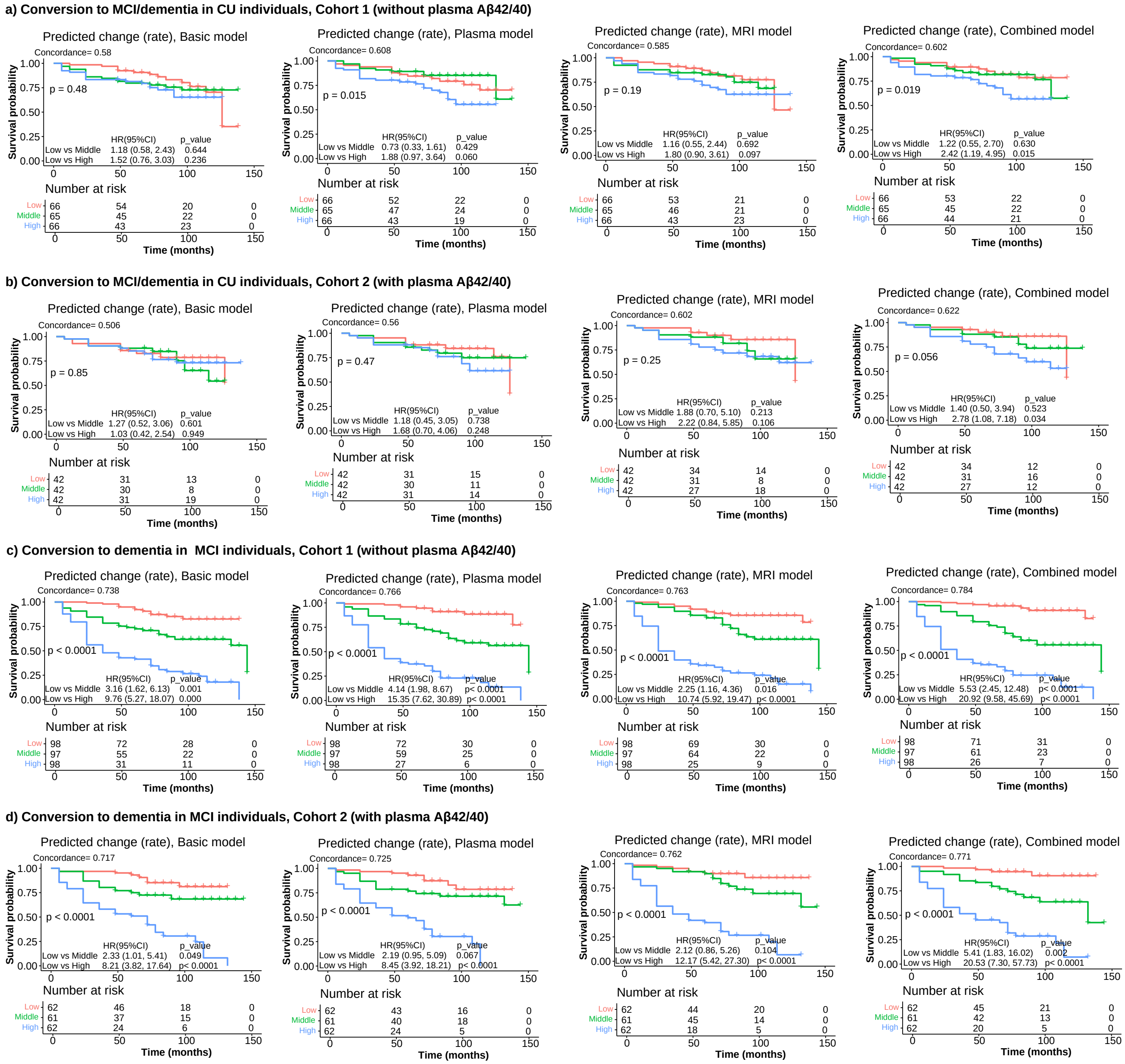


Figure S4: Kaplan-Meier survival curves for conversion to MCI/AD in CU and MCI individuals: The predicted rate of cognitive decline in ADNI-LAN score was divided into 3 tertiles (low, middle, high). Vertical tick marks on lines indicate times at which the individual was censored. p-values are for log-rank tests among the tertiles. Plasma model: Demographics, APOE4, Composite cognitive scores, plasma biomarkers. MRI model: Demographics, APOE4, Composite cognitive scores, MRI data. Combined model: Demographics, APOE4, Composite cognitive scores, plasma biomarkers, MRI data.

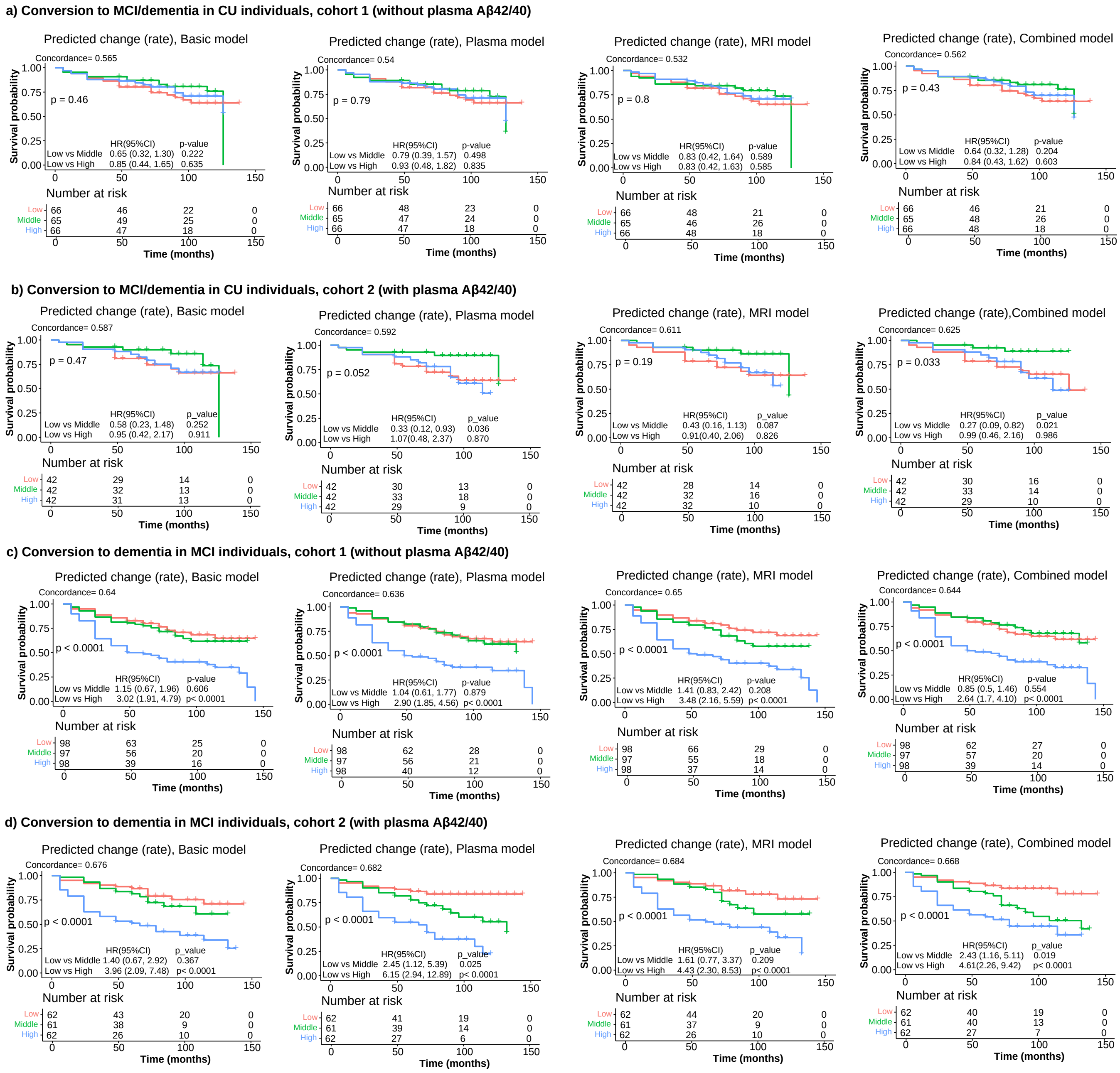


Figure S5: Kaplan-Meier survival curves for conversion to MCI/AD in CU and MCI individuals: The predicted rate of cognitive decline in ADNI-VS score was divided into 3 tertiles (low, middle, high). Vertical tick marks on lines indicate times at which the individual was censored. p-values are for log-rank tests among the tertiles. Plasma model: Demographics, APOE4, Composite cognitive scores, plasma biomarkers. MRI model: Demographics, APOE4, Composite cognitive scores, MRI data. Combined model: Demographics, APOE4, Composite cognitive scores, plasma biomarkers, MRI data.
